# Autoantibodies to nuclear valosin-containing protein-like protein: identification and characterization of systemic sclerosis-related anti-nucleolar antibodies utilizing *in vitro* human proteome

**DOI:** 10.1101/2023.07.16.23292097

**Authors:** Kazuki M Matsuda, Hirohito Kotani, Kei Yamaguchi, Chihiro Ono, Taishi Okumura, Koji Ogawa, Ayako Miya, Ayaka Sato, Rikako Uchino, Murakami Yumi, Hiroshi Matsunaka, Masanori Kono, Yuta Norimatsu, Teruyoshi Hisamoto, Ruriko Kawanabe, Ai Kuzumi, Takemichi Fukasawa, Asako Yoshizaki-Ogawa, Tomohisa Okamura, Hirofumi Shoda, Keishi Fujio, Takashi Matsushita, Naoki Goshima, Shinichi Sato, Ayumi Yoshizaki

## Abstract

**Objectives:** To identify and characterize undescribed systemic sclerosis (SSc)-related autoantibodies targeting nucleolar antigens and to assess their clinical significance.

**Methods:** We conducted proteome-wide autoantibody screening (PWAS) against serum samples from SSc patients with nucleolar patterned anti-nuclear antibodies (NUC-ANAs) of specific antibodies (Abs) unknown, utilizing wet protein arrays fabricated from *in vitro* human proteome. Controls included SSc patients with already-known SSc-related autoantibodies, patients with other connective tissue diseases, and healthy subjects. The selection of nucleolar antigens was performed by database search in the Human Protein Atlas. The Presence of autoantibodies was certified by immunoblots, indirect immunofluorescence assays, and immunoprecipitations. Clinical assessment was conducted by retrospective review of electric medical records.

**Results:** PWAS identified autoantibodies targeting nuclear valosin-containing protein-like (NVL), DIM1 rRNA methyltransferase ribosome maturation factor (DIMT1), and telomeric repeat binding factor 1 (TERF1) as candidates. Additional measurements in disease controls revealed that only anti-NVL Abs are exclusively detected in SSc. Detection of anti-NVL Abs was reproduced by conventional assays. Anti-NVL Ab-positive cases were characterized by significantly low prevalence of diffuse skin sclerosis and interstitial lung disease, compared to SSc cases with NUC-ANAs other than anti-NVL Abs, such as anti-U3-RNP and anti-Th/To Abs.

**Conclusions:** Anti-NVL Ab is an SSc-related autoantibody associated with a unique combination of clinical features, including limited skin sclerosis and lack of lung involvement.

## Introduction

Systemic sclerosis (SSc) is a multi-organ connective tissue disease characterized by a triad of tissue fibrosis, vasculopathy, and immune system abnormality.(1) Although the most part of the pathophysiology remains unclear, it is considered that autoimmunity plays an important role in the pathogenesis. Anti-nuclear antibodies (ANAs) are detected in more than 95% of the patients,(2) which highlights the autoimmune aspect of the disease. Several ANAs are known to be SSc-specific, and moreover, closely associated with distinct clinical subsets.(3) For instance, anti-topoisomerase I antibodies (ATA) and anti-RNA polymerase III antibodies (ARA) have close association with diffuse skin sclerosis, while limited skin involvement is dominant among those with anti-centromere antibodies (ACA). Meanwhile, interstitial lung disease (ILD), the leading cause of death in SSc,(4) is common among ATA-positive patients, whereas the prevalence of ILD is low in ARA or ACA-positive cases. Therefore, detection of such SSc-related antibodies (Abs) is vital in the diagnosis, prediction of the disease course, and determining the therapeutic strategies.

The approach to identify SSc-related Abs in daily clinical practice consists of two steps. In the first step, clinicians submit the patient’s serum for primary ANA screening by indirect immunofluorescence (IIF).(5) The results are returned as combination of antibody titers and their staining patterns, such as homogenous, speckled, discrete speckled, and nucleolar patterns. While the discrete speckled type is almost identical to the presence of ACA, most other staining patterns cannot be narrowed down to specific SSc-related Abs. Thus, in the second step, we specifically measure several autoantibodies expected from the ANA staining pattern, generally utilizing immunologic tests such as enzyme-linked immunosorbent assays (ELISA,) double immunodiffusion assays, and immunoprecipitation (IP) assays.(6) However, even in accordance with this strategy, specific antibodies cannot be identified in all cases. Such cases of specific antibodies unknown are often difficult to diagnose and to manage, resulting in a significant burden for both the patient and the healthcare providers.

In particular, detection of nucleolar patterned-ANA (NUC-ANA) in IIF indicates some SSc-related autoantibodies with outstanding clinical importance, such as anti-U3-RNP, anti-Th/To, and anti-PM-Scl Abs. The presence of NUC-ANAs in SSc is generally thought to be indicative of a more severe disease course. Indeed, anti-U3-RNP Ab is known to be closely associated with severe visceral involvement, including cardiac, renal, and gastrointestinal involvement;(7) anti-Th/To Ab with severe peripheral vasculopathy and pulmonary hypertension;(8) and anti-PM-Scl Ab with myositis overlap.(9) Meanwhile, not a few cases with NUC-ANA fail to identify specific SSc-related antibodies, some of whom experience a mild disease course. Hence, researchers have expected the existence of an undescribed NUC-ANA associated with minor disease manifestations.

Herein, we performed a proteome-wide autoantibody screening (PWAS) using wet protein arrays (WPAs) fabricated from our *in vitro* human proteome,(10)(11)(12) to discover novel SSc-related Abs in serum samples from SSc patients with NUC-ANAs of specific antibodies unknown. As a result, we identified autoantibodies targeting nuclear valosin-containing protein-like (NVL) protein uniquely in SSc. The presence of anti-NVL Abs was certified by conventional studies, including immunoblots, IIF assays, and IP assays. Further clinical assessment revealed that anti-NVL is associated with limited skin sclerosis and lack of ILD.

## Methods

### Patients and controls

We consecutively enrolled Japanese patients with SSc visited our clinic between February 2019 and June 2021. Of these, those who showed a nucleolar pattern in ANA screening by IIF with negative screening results in SSc-related Abs, including ATA, ACA, ARA, anti-U3-RNP Ab, anti-Th/To Ab, and anti-PM-Scl Ab, were selected, framed as the “Tokyo cohort (n = 4).” Additional SSc patients were obtained from Kanazawa University according to the same protocol between August 2002 and May 2019, defined as the “Kanazawa cohort (n = 8).” Patients with anti-U3-RNP or Th/To Abs were also pooled for clinical comparison. SSc patients with ATA, ARA, or ACA, as well as patients with dermatomyositis (DM) or systemic lupus erythematosus (SLE) who visited The University of Tokyo Hospital during the same period were randomly recruited as disease controls. All the patients fulfilled the relevant classification criteria.(13)(14)(15) Healthy controls (HCs) were collected during annual checkups from those without any medical history. Basic information about the participants is described in **Supplementary Table 1**. This study has been approved by The University of Tokyo Ethical Committee (Approval number 0695). Written informed consent has been obtained from all the participants.

### Clinical assessments

Clinical and laboratory information was collected by retrospective review of electric medical records from the closest time point from the date of serum collection. SSc patients were classified into diffuse cutaneous SSc (dcSSc) and limited cutaneous SSc (lcSSc) based on LeRoy’s classification criteria, as well as SSc in overlap showing features of other collagen diseases.(16) The definitions for organ system involvement attributable to SSc are as described previously.(17)

### Autoantibody measurement by WPAs

PWAS for serum samples were performed utilizing WPAs as previously described.(12) First, proteins with GST and FLAG tags added on their N-terminus were expressed *in vitro* from 13,355 entry cDNA clones covering approximately 90% of the human transcriptome (**Supplementary Table 2**),(10) utilizing a wheat germ cell-free system.(18) Second, proteins were plotted onto glass slides in an array format.(11) Human serum was diluted by 3:1000, added to WPAs, and reacted for 1 hour at room temperature. Next, the WPA was washed, and goat anti-Human IgG (H+L) Alexa Flour 647 conjugate (Thermo Fisher Scientific, San Jose, CA, USA) diluted 1000-fold was added to the WPA and reacted for 1 hour at room temperature. Finally, the WPA was washed, air-dried, and fluorescent images were acquired using a fluorescence imager Typhoon FLA 9500 (Cytiva, Marlborough, MA, USA). Fluorescence images were analyzed to identify or quantify autoantibodies according to the formula shown below. Serum autoantibody level higher than 1.5625 (1/64 of positive control) was framed as autoantibody positivity.

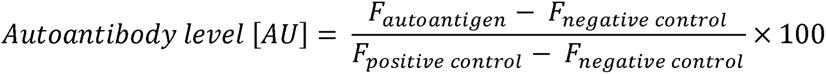

*AU*: arbitrary unit

*F _autoantigen_*: fluorescent intensity of autoantigen spot

*F _negative control_*: fluorescent intensity of negative control spot

*F _positive control_*: fluorescent intensity of positive control spot

### Database search

Subcellular location of the proteins targeted by autoantibodies detected in PWAS for serum samples from SSc patients with NUC-ANAs of specific autoantibodies unknown was searched in the Human Protein Atlas,(19) utilizing Metascape, a gene annotation & analysis resource.(20)

### Immunoblots

Proteins were extracted from HEK293 cells cultured in DMEM (Thermo Fisher Scientific) at 37□°C with 5% CO_2_, using RIPA lysis buffer (Santa Cruz Biotechnology, Santa Cruz, CA, USA) according to the manufacturer’s manual. Extracted proteins were separated by sodium dodecyl sulfate-polyacrylamide gel electrophoresis (SDS-PAGE) and transferred to polyvinylidene difluoride (PVDF) membranes (Millipore, Bedford, MA, USA). Membranes were blocked with 5% non-fat milk and 1% bovine serum albumin (Iwai Chemicals, Tokyo, Japan) in Tris-buffered saline /0.05% Tween 20 (Cell Signaling Technology, Danvers, MA, USA) for 30 minutes at room temperature and incubated overnight at 4□°C with human sera diluted 250-fold or rabbit anti-human NVL Ab (Sigma Aldrich, St. Louis, MZ, USA) diluted 1000-fold, followed by incubation with horseradish peroxidase-conjugated secondary antibodies (Cappel Research Reagents, Cochranville, PA, USA) for one hour at room temperature. Proteins were visualized using Chemi-Lumi One Ultra (Nacalai Tesque, Kyoto, Japan).

### IIF assays

ANA was detected by IIF performed on Hep-2 cell slides as the substitute, combined with fluorescein isothiocyanate-conjugated anti-human IgG (Medical & Biological Laboratories, Tokyo, Japan). In some experiments, rabbit anti-human NVL Ab (Sigma Aldrich) diluted at 1:50 was used in combination with Alexa Fluor 594-conjugated anti-rabbit IgG (Thermo Fisher Scientific). The specimens were observed with a fluorescence microscope Bio Zero BZ-X810 (Keyence, Osaka, Japan).

### IP assays

IP assays were performed using extract of a leukemia cell line K562, as previously described.(21) Briefly, 10 μL of patients’ sera were mixed with protein A-Sepharose CL-4B (Pharmacia Biotech). Antibody-coated Sepharose beads were incubated with ^35^S-methionine-labeled K562 cellular extracts, and immunoprecipitated materials were subjected to SDS-PAGE. Radiolabeled polypeptide components were analyzed by autoradiography.

### Epitope mapping

We synthesized full-length and truncated forms of NVL protein from our entry clone (NCBI Reference Sequence: NM_001243146.2) with GST and FLAG tags added on their N-terminus using a wheat germ cell-free system. We utilized Polydot in Sequence Analysis Software (EBBOSS, Hinxton, Cambridge, UK) to generate dot plots for each sequence to compare the amino acid sequence homology of the truncated forms of NVL protein. The synthesized proteins were confirmed to match the target size by SDS-PAGE and Coomassie Brilliant Blue staining and attached to 96-well plates coated with glutathione by its affinity with GST tags. The antigen-coated plates were reacted with Alexa Fluor 647-conjugated anti-FLAG Ab (MBL, Tokyo, Japan) diluted by 1:3000 for 1 hour at room temperature. After washing the plate, fluorescence was measured by a fluorescence imager (Typhoon FLA 9500). Next, the plates were washed again and treated with human serum diluted by 1:150 for 1 hour at room temperature, and subsequently with Alexa Fluor 647-conjugated goat anti-human IgG (H+L) Ab (Thermo Fisher Scientific). After washing the plates, fluorescence was measured by a fluorescence imager (Typhoon FLA 9500). Fluorescence measurements obtained from each well were corrected by the following formula.

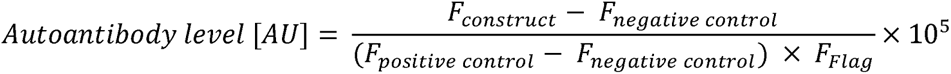

*AU*: arbitrary unit

*F _construct_*: fluorescent intensity of a spot coated by each construct

*F _negative control_*: fluorescent intensity of a null spot as a negative control

*F _positive control_*: fluorescent intensity of a spot coated by IgG as a positive control

### Statistical analysis

Statistical analyses were performed by Mann-Whitney’s *U* test or Kruskal Wallis test for the comparison of continuous variables and Fisher’s exact test for comparing categorical variables. *P* values lower than 0.05 was considered to be statistically significant. Spearman’s r was calculated to produce the correlation matrix. All the analyses were performed using Prism 9 (GraphPad Software, CA, USA).

## Results

### Detection of three novel autoantibody candidates exhibiting NUC-ANAs in IIF by WPAs

We recruited four SSc patients with NUC-ANA positivity in the Tokyo cohort for PWAS. Among all the human proteins displayed on the WPAs, 292 items reacted with more than two serum samples (**Supplementary Table 3**). Of these, 25 proteins were found to be localized in the nucleoli using the Human Protein Atlas. After excluding proteins reacting with the healthy controls (n = 46), three autoantibodies, including anti-NVL Abs, finally remained as candidates for SSc-related anti-nucleolar Abs (**Fig. 1A**).

**Figure 1.**
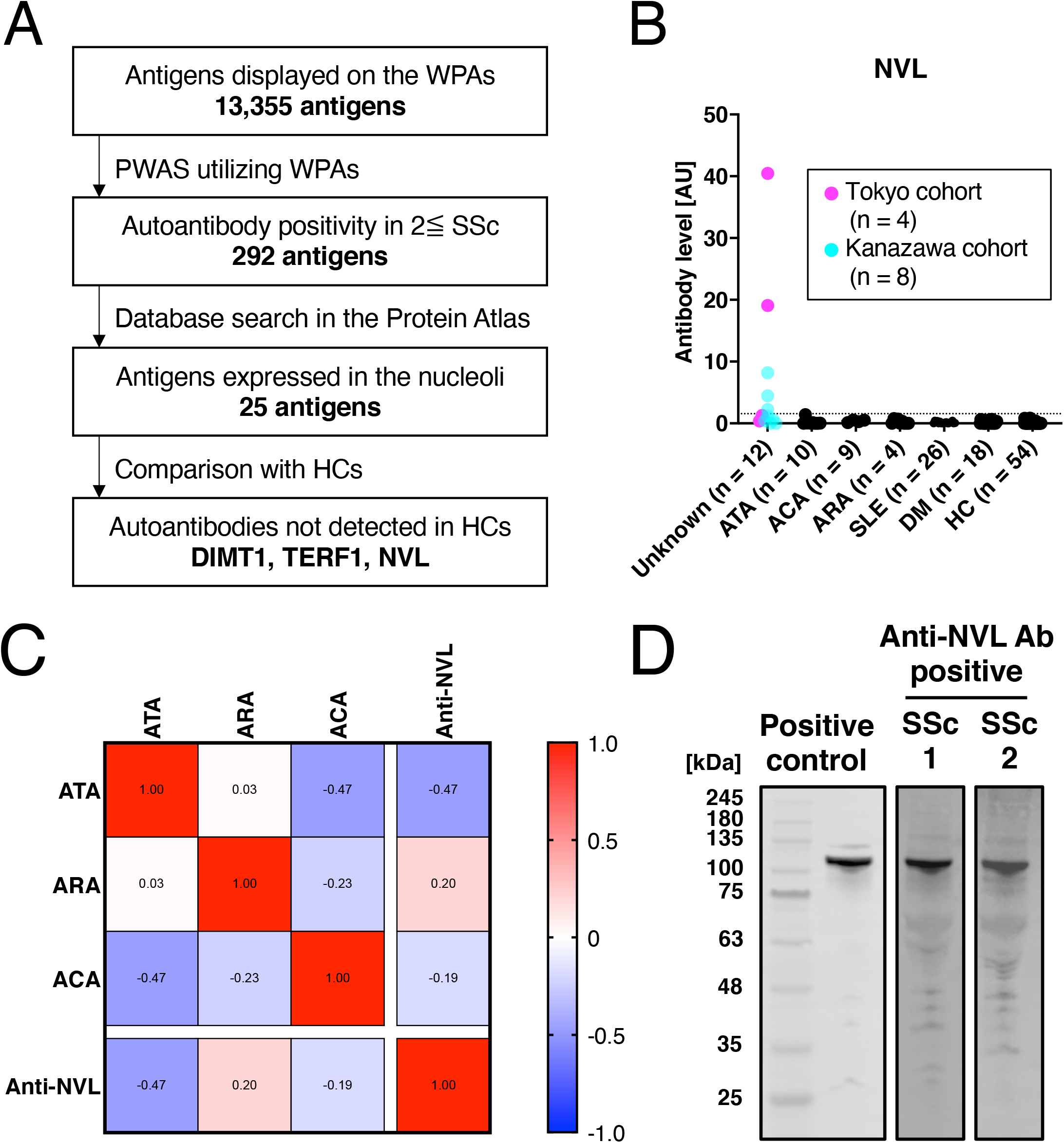
Identification of anti-NVL Abs by PWAS in SSc. **(A)** Flow-chart of autoantibody selection. **(B)** Serum levels of anti-NVL Abs in SSc patients with NUC-ANAs of specific autoantibodies unknown (Unknown), ATA positive SSc patients, ACA positive SSc patients, ARA positive SSc patients, DM patients, SLE patients, and HCs. **(C)** Correlation matrix of ATA, ARA, ACA, and anti-NVL Abs in the sera of SSc patients. The value in each box and the color bar indicates Spearman’s r. **(D)** Immunoblots for detecting autoantibodies to NVL in two SSc cases in the Tokyo cohort. The rabbit anti-human NVL Ab was used as a positive control.

### Anti-NVL antibody is specific to SSc

PWAS was additionally performed for eight members of the Kanazawa cohort, SSc patients with ATA, ACA, and ARA, DM patients, and SLE patients. As a result, positivity to anti-NVL Ab was specifically detected in the Tokyo and Kanazawa cohorts (**Fig. 1B**), whereas higher serum levels of the other two candidates were observed in DM than in SSc (**Supplementary Fig. 1**). Moreover, correlation analysis subjecting SSc patients demonstrated low association among the serum levels of ATA, ACA, ARA, and anti-NVL Abs (**Fig. 1C**), suggesting the uniqueness of anti-NVL Ab in comparison with the already-known SSc-related Abs.

### Certification of anti-NVL Ab-positivity by conventional assays

We subsequently carried out immunoblots using extracts of HEK293 cells, which showed that anti-NVL Ab-positive sera from the Tokyo cohort and rabbit anti-human NVL IgG illustrate bands at the same molecular weight (**Fig. 1D**). ANA assays using Hep-2 cells also demonstrated similar nucleolar patterns by anti-NVL Ab-positive sera in the Tokyo cohort and rabbit anti-human NVL IgG (**Fig. 2**). Furthermore, IP assays for the Kanazawa cohort using extracts of K562 cells demonstrated bands nearby the markers indicating 97 kDa with anti-NVL Ab-positive sera, while no such bands appeared with anti-NVL Ab-negative sera (**Fig. 3**). Collectively, the presence of anti-NVL Abs in the sera of SSc patients were replicated by methodologies other than PWAS utilizing WPAs, subjecting multiple human cell lines.

**Figure 2.**
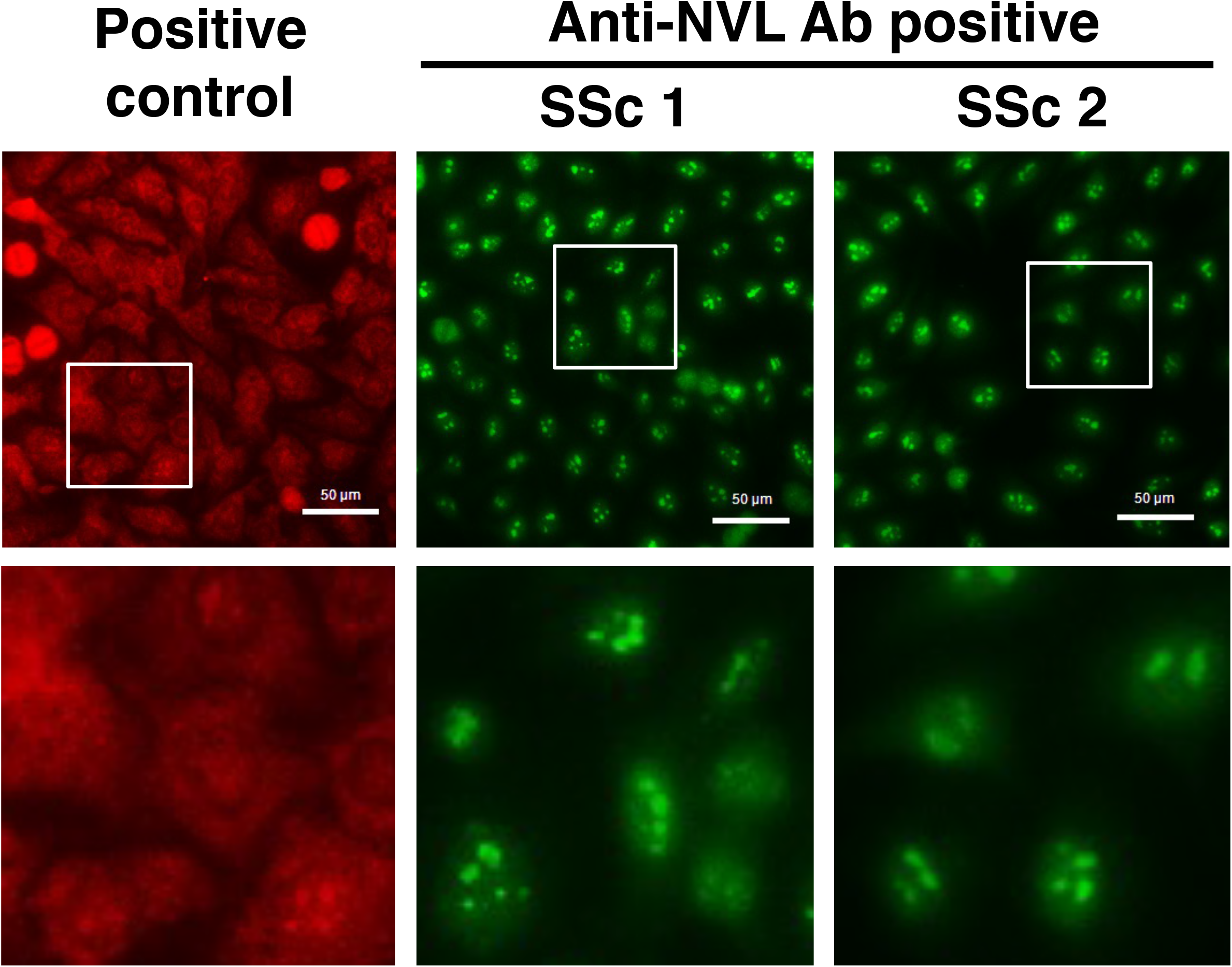
Detection of anti-NVL Abs by IIF assays. IIF staining patterns on Hep-2 cells produced by the sera of two SSc cases in the Tokyo cohort, demonstrating nucleolar patterns. The rabbit anti-human NVL Ab was used as a positive control. High-magnification views of the white framed area are displayed on the bottom row.

**Figure 3.**
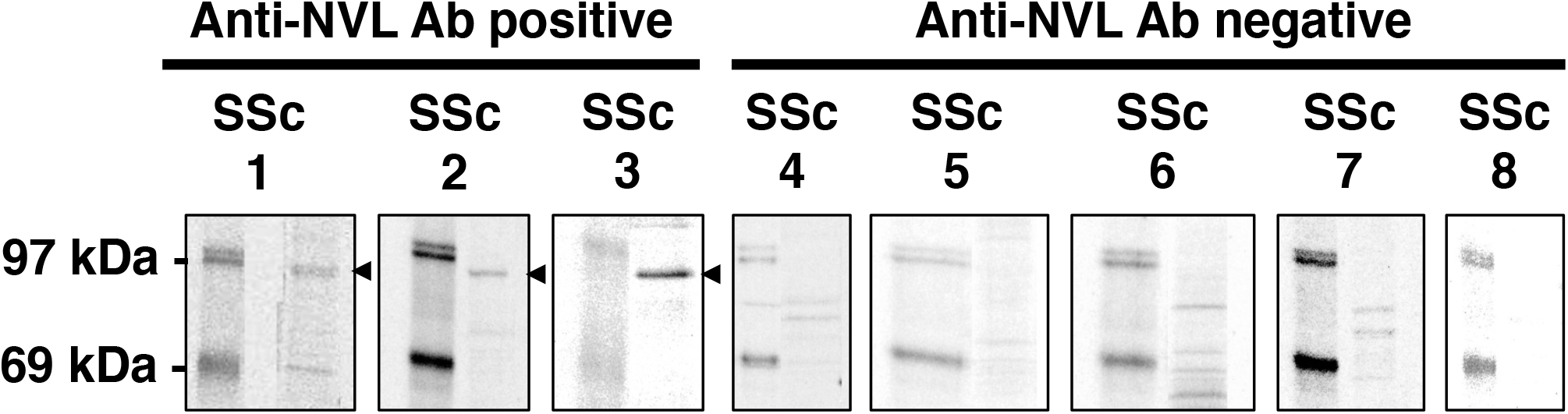
Detection of anti-NVL Abs by IP assays. Immunoprecipitants from ^35^S-methionine-labeled K562 cellular extracts were subjected to SDS-PAGE, followed by autoradiography. Each column represents a serum sample from SSc cases in the Kanazawa cohort. Within each column, the left lane = molecular weight markers, and the right lane = immunoprecipitants. **Arrowheads** show the bands indicative for the presence of anti-NVL Abs.

### Epitope mapping

We conducted an epitope mapping of anti-NVL antibodies by creating truncated forms of NVL based on their domains, as illustrated in **Fig. 4A**. We discovered only minor homology between NVL_D and NVL_A, as well as NVL_B, but none with the other variants (**Fig. 4B**). Epitope mapping was performed on sera from four SSc patients with NUC-ANAs of specific autoantibodies unknown in the Tokyo cohort: two were positive for anti-NVL antibodies and two were negative, with all displaying nucleolar patterns in ANA assays. Interestingly, both anti-NVL antibody-positive sera identified autoantibody levels against NVL_A, NVL_C, NVL_D, and NVL_E, but not against NVL_B (**Fig. 4C**). Moreover, when we conducted the epitope mapping with shorter truncated forms, autoantibody levels were consistently detected against NVL_A1 and NVL_E2. However, the detection was weaker and less common against the shorter truncated forms of NVL_C and NVL_D. These findings suggested that anti-NVL antibodies target distinct epitopes located in various regions of the NVL protein.

**Figure 4.**
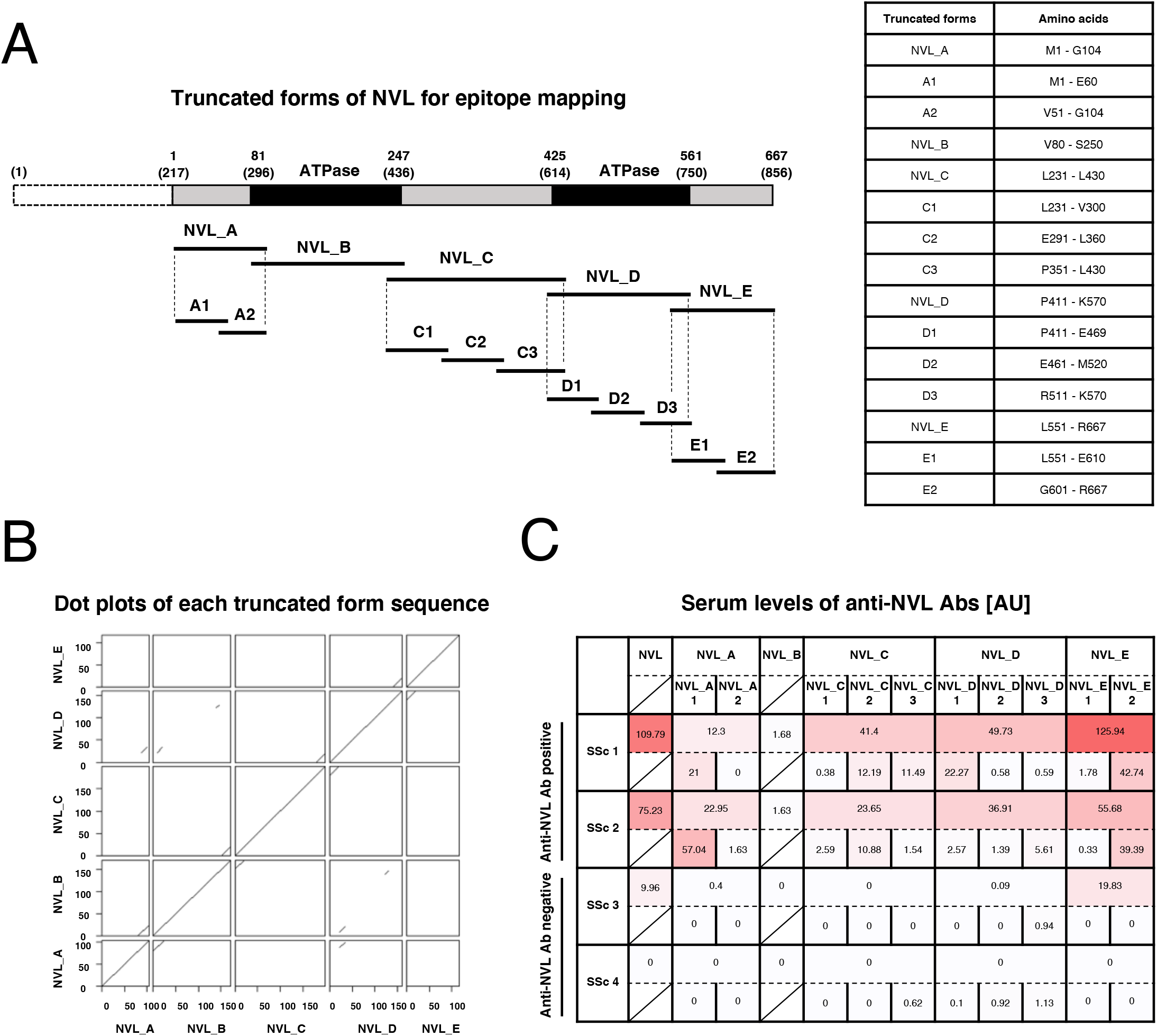
Epitope mapping. **(A)** The design of truncated forms of NVL protein synthesized for epitope mapping. The black regions indicate the AAA+ ATPase domain (ATPase). Numbers indicate amino acids from the initial methionine. Numbers in parentheses indicate the corresponding positions of amino acids within the canonical sequence. **(B)** Dot plots of the amino acid sequence of the truncated constructs: NVL_A-E. The numbers on the vertical and horizontal axes correspond to the number of amino acid residues from the N-terminus in each fragment. **(C)** Heatmap illustrating the autoantibody levels against full-length or each truncated form of NVL among the SSc patients in the Tokyo cohort. AU: arbitrary unit.

### Clinical features of SSc associated with anti-NVL Abs

We divided SSc patients in the Tokyo and Kanazawa cohort into two groups according to the positivity of anti-NVL Abs and compared clinical characteristics (**Table 1**). Since the number of SSc patients was small in each cohort, statistical analysis was conducted by combining the two cohorts into one. Notably, diffuse skin sclerosis was less common in patients with anti-NVL Abs compared to the others (P = 0.023). In terms of organ involvement, none of the patients with anti-NVL Ab-positivity had ILD, while ILD was frequently observed in those without anti-NVL Ab (P = 0.028). To further examine potential differences in the clinical significance of anti-NVL Abs and already-known SSc-related anti-nucleolar Abs, clinical features were compared among SSc patients for whom anti-NVL, Th/To, and U3-RNP Abs were identified (**Table 2**). For this analysis, SSc patients in the Tokyo and Kanazawa cohorts were combined. In line with the association analysis shown above, proportion of patients with lcSSc was the highest in those with anti-NVL Abs among the three groups (P = 0.012). Additionally, the prevalence of ILD was the lowest among anti-NVL Ab-positive patients (P = 0.028). Collectively, anti-NVL Abs seemed to be associated with relatively minor skin and lung fibrosis among SSc patients with nucleolar patterned ANAs.

**Table 1.**
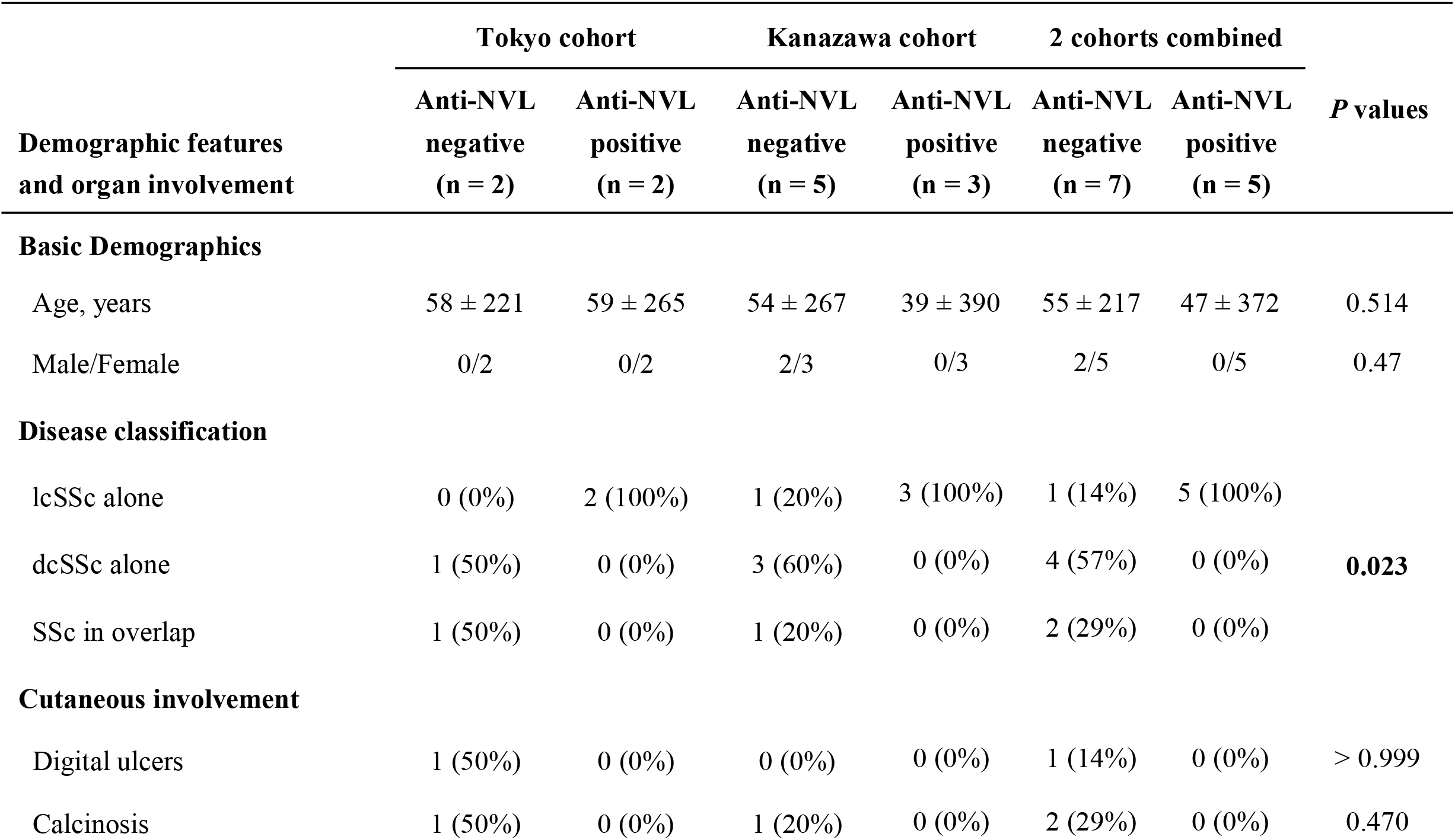
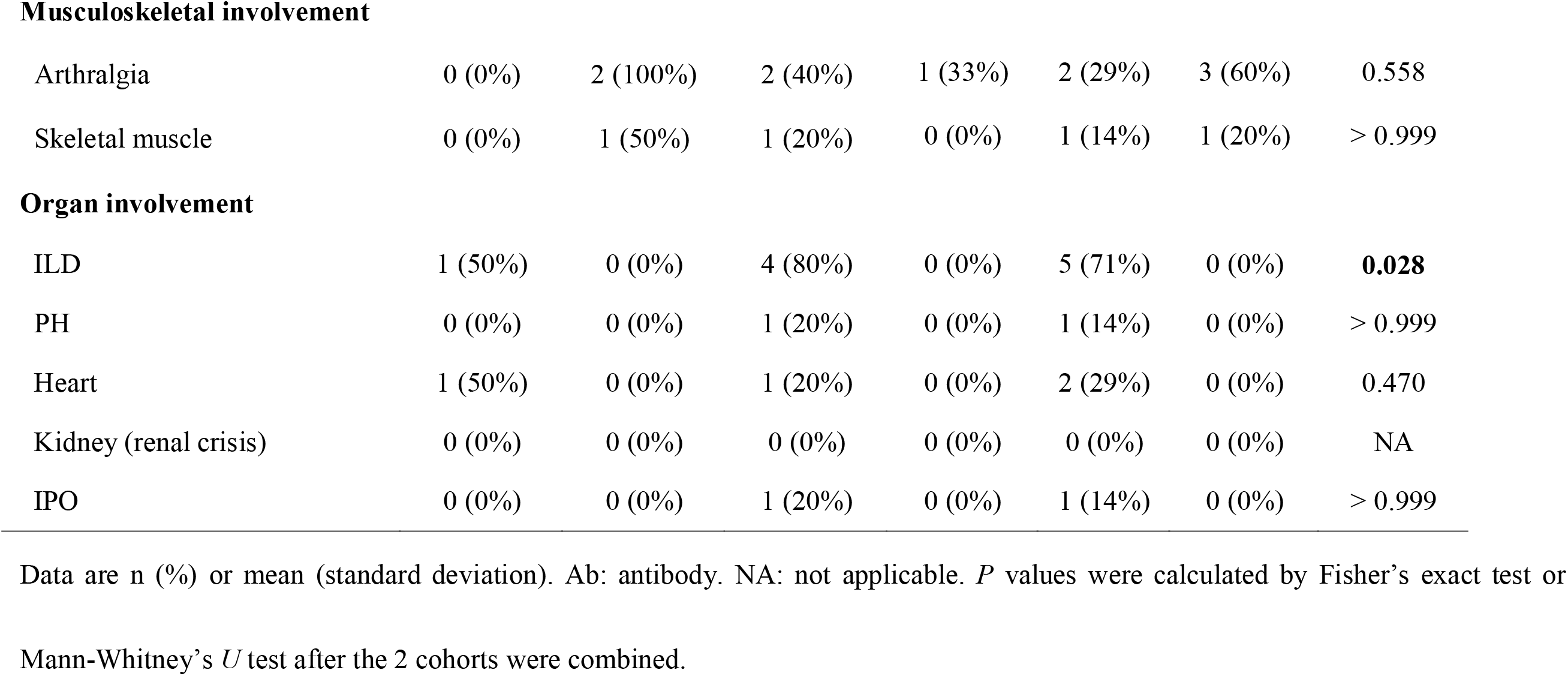
Clinical profiles of SSc patients with or without anti-NVL antibody in 2 independent Japanese cohorts.

|  | Tokyo cohort |  | Kanazawa cohort |  | 2 cohorts combined |  | *P* values |
| --- | --- | --- | --- | --- | --- | --- | --- |
|  | Anti-NVL<br>negative<br>(n = 2) | Anti-NVL<br>positive<br>(n = 2) | Anti-NVL<br>negative<br>(n = 5) | Anti-NVL<br>positive<br>(n = 3) | Anti-NVL<br>negative<br>(n = 7) | Anti-NVL<br>positive<br>(n = 5) |  |
| Demographic features and organ involvement |  |  |  |  |  |  |  |
| Basic Demographics |  |  |  |  |  |  |  |
| Age, years | 58 ± 221 | 59 ± 265 | 54 ± 267 | 39 ± 390 | 55 ± 217 | 47 ± 372 | 0.514 |
| Male/Female | 0/2 | 0/2 | 2/3 | 0/3 | 2/5 | 0/5 | 0.47 |
| Disease classification |  |  |  |  |  |  |  |
| lcSSc alone | 0 (0%) | 2 (100%) | 1 (20%) | 3 (100%) | 1 (14%) | 5 (100%) | 0.023 |
| dcSSc alone | 1 (50%) | 0 (0%) | 3 (60%) | 0 (0%) | 4 (57%) | 0 (0%) |  |
| SSc in overlap | 1 (50%) | 0 (0%) | 1 (20%) | 0 (0%) | 2 (29%) | 0 (0%) |  |
| Cutaneous involvement |  |  |  |  |  |  |  |
| Digital ulcers | 1 (50%) | 0 (0%) | 0 (0%) | 0 (0%) | 1 (14%) | 0 (0%) | > 0.999 |
| Calcinosis | 1 (50%) | 0 (0%) | 1 (20%) | 0 (0%) | 2 (29%) | 0 (0%) | 0.470 |
| **Musculoskeletal involvement** |  |  |  |  |  |  |  |
| Arthralgia | 0 (0%) | 2 (100%) | 2 (40%) | 1 (33%) | 2 (29%) | 3 (60%) | 0.558 |
| Skeletal muscle | 0 (0%) | 1 (50%) | 1 (20%) | 0 (0%) | 1 (14%) | 1 (20%) | > 0.999 |
| **Organ involvement** |  |  |  |  |  |  |  |
| ILD | 1 (50%) | 0 (0%) | 4 (80%) | 0 (0%) | 5 (71%) | 0 (0%) | **0.028** |
| PH | 0 (0%) | 0 (0%) | 1 (20%) | 0 (0%) | 1 (14%) | 0 (0%) | > 0.999 |
| Heart | 1 (50%) | 0 (0%) | 1 (20%) | 0 (0%) | 2 (29%) | 0 (0%) | 0.470 |
| Kidney (renal crisis) | 0 (0%) | 0 (0%) | 0 (0%) | 0 (0%) | 0 (0%) | 0 (0%) | NA |
| IPO | 0 (0%) | 0 (0%) | 1 (20%) | 0 (0%) | 1 (14%) | 0 (0%) | > 0.999 |

Data are n (%) or mean (standard deviation). Ab: antibody. NA: not applicable. *P* values were calculated by Fisher's exact test or
Mann-Whitney's *U* test after the 2 cohorts were combined.

**Table 2.** Comparison of clinical profiles in SSc patients with 3 anti-nucleolar autoantibodies.

| **Demographic features**<br>**and organ involvement** | **Anti-NVL Ab positive**<br>**(n = 5)** | **Anti-Th/To Ab positive**<br>**(n = 3)** | **Anti-U3-RNP Ab positive**<br>**(n = 7)** | **Overall**<br>**P values** |
| --- | --- | --- | --- | --- |
| **Basic Demographics** |  |  |  |  |
| Age, years | 47 ± 372 | 64 ± 49 | 66 ± 313 |  |
| Male/Female | 0/5 | 1/2 | 2/5 | 0.385 |
| **Disease classification** |  |  |  |  |
| lcSSc alone | 5 (100%) | 0 (0%) | 1 (14%) | **0.012** |
| dcSSc alone | 0 (0%) | 2 (67%) | 4 (57%) |  |
| SSc in overlap | 0 (0%) | 1 (33%) | 2 (29%) |  |
| **Cutaneous involvement** |  |  |  |  |
| Digital ulcers | 0 (0%) | 1 (33%) | 3 (43%) | 0.333 |
| Calcinosis | 0 (0%) | 0 (0%) | 1 (14%) | > 0.999 |
| **Musculoskeletal involvement** |  |  |  |  |
| Arthralgia | 3 (60%) | 2 (67%) | 2 (29%) | 0.511 |
| Skeletal muscle | 1 (20%) | 1 (33%) | 2 (29%) | > 0.999 |
| **Organ involvement** |  |  |  |  |
| ILD | 0 (0%) | 1 (33%) | 5 (71%) | **0.028** |
| PH | 0 (0%) | 0 (0%) | 3 (43%) | 0.246 |
| Heart | 0 (0%) | 0 (0%) | 2 (29%) | 0.667 |
| Kidney (renal crisis) | 0 (0%) | 0 (0%) | 1 (14%) | > 0.999 |
| IPO | 0 (0%) | 0 (0%) | 3 (43%) | 0.246 |

Data are n (%) or mean (standard deviation). Ab: antibody. NA: not applicable. *P* values were calculated by Fisher's exact test or Kruskal-Wallis test.

## Discussion

In this study, we identified autoantibodies to NVL in a small group of SSc patients with NUC-ANA by PWAS utilizing WPAs (**Fig. 1A**). Presence of anti-NVL Abs was specific to SSc (**Fig. 1B**), and moreover, seemed to be independent from already-known SSc-related Abs such as ATA, ACA, and ARA (**Fig. 1C**). Anti-NVL Abs were also detectable by conventional techniques including immunoblots (**Fig. 1D**), IIF assays (**Fig. 2**), and IP assays, using different human cell lines as substrates (**Fig. 3**). Although Vulsteke et al. had already indicated the possibility of NVL as a novel SSc-related Ab by protein immunoprecipitation combined with gel-free liquid chromatography-tandem mass spectrometry,(22) we additionally found that the major epitope places in both the N-terminal and the C-terminal regions of the protein (**Fig. 4**), and moreover, demonstrated association among anti-NVL Abs, limited skin sclerosis, and lack of ILD among SSc patients with NUC-ANA by clinical assessments (**Tables 1 and 2**). Given together, we suggest anti-NVL Ab should be added to the list of SSc-related autoantibodies, which would provide us with great clinical benefit.

NVL is a protein with similar structure to valosin-containing protein, which locates in the nucleoli of the cells.(23) NVL belongs to a group of proteins called AAA+ ATPases, and plays a role in RNA metabolism and processing, especially in the splicing of pre-ribosomal RNA.(24) NVL has also been linked to the regulation of the assembly of the telomerase holoenzyme and effecting of telomerase activity.(25) Involvement in RNA processing and genome structure maintenance is common with several nuclear antigens targeted by already-known SSc-related Abs, such as RuvBL1/2, Ku, and PM-Scl. However, the role of these autoantibodies in the pathogenesis is questionable, because Abs cannot usually access the nuclear antigens across the plasma and nuclear membranes *in vivo*,(26) and moreover, disturbance of RNA processing or DNA repair would be fatal for the organism and is less likely to result in the manifestations of SSc as clinical phenotypes. Pathogenicity of anti-NVL Abs should be further investigated by additional experiments, i.e., immunization of animals with the relevant antigen.(27) Alternatively, it would be insightful to examine the correlation between serum concentrations of anti-NVL Ab and disease activity or response to therapy, as observed in diseases with established autoantibody pathogenicity, such as autoimmune bullous diseases.(28)

Our epitope mapping analysis demonstrated that the primary epitope targeted by autoantibodies against NVL is located in both the N-terminal and C-terminal regions of the protein, as depicted in **Fig 4C**. Notably, no significant amino acid sequence nor functional similarities were observed between these two regions (**Fig 4A and 4B**). Interestingly, the presence of multiple epitopes lacking homology to one another is a characteristic shared by other SSc-associated autoantibodies, including ATA(29). These findings suggest the polyclonal nature of autoantibody responses in SSc; however, further confirmation is warranted. Furthermore, we can propose a hypothesis that intramolecular epitope spreading may be associated with the disease progression of SSc.(30) However, the validation of this hypothesis requires longitudinal data to support it.(31)

Autoantibodies to telomeric repeat binding factor 1 (TERF1), one of the two candidates for SSc-related autoantibodies other than anti-NVL Ab revealed by our PWAS (**Fig 1A and Supplementary Fig 1**), have also been found in a subset of patients with SSc and idiopathic pulmonary fibrosis by IP assays and ELISA.(32) Their investigation demonstrated the association between anti-TERF1 Abs and short lymphocyte telomere length and pulmonary fibrosis. In addition, another group has reported detection of anti-TERF1 Abs in the sera of SSc patients by IP-MS.(22) Our study reproduced their insights in terms of detecting anti-TERF1 Abs in serum samples utilizing our WPA, although not exclusively in SSc. Further examination of the serum positivity to autoantibodies targeting telomere-related proteins, including NVL and TERF1, and their association with clinical manifestations or telomere length in our Japanese cohort would be an attractive research subject.

Our PWAS method utilizing WPAs based on *in vitro* human proteome has multiple strengths compared to previous methodologies. First, combination of the comprehensiveness of our cDNA library and the high-throughput capability of the *in vitro* protein synthesis technology using the wheat germ cell-free system has achieved a high coverage rate at 90% of the human transcriptome.(10)(18) Second, the three-dimensional structure of the antigen can be assayed in a condition close to its physiological state by completing the entire assay process in a non-dried environment.(10) Third, high-density protein plotting technique made it possible to display more than 13,000 antigens on relatively small glass slides, resulting in achieving high-dimensional information even from a small sample amount.(11) Fourth, there is no risk of radiation exposure because no radioisotopes are used, and moreover, the entire assay takes only about six hours.(12) These features indicate that our technology is advantageous not only in exploratory PWAS, but also in the practical application of individual measurement or panel testing for autoantibodies.(12)(33)

The present study has several limitations. First, the cutoff values for positivity in PWAS were uniform and not optimized for each item, which might be causing clinically important autoantibodies to be missed. An alternative would be to set individual thresholds (e.g., mean + 3 standard deviations) for each antigen based on the measurement results in HCs. Second, the number of patients included in the clinical assessment was limited mainly due to the rarity of anti-NVL antibodies and the fact that this study only included NUC-ANA positive cases. Surveillance of anti-NVL Abs targeting a larger SSc cohort is desirable in the future. Fourth, the data in this study was limited to two Japanese cohorts and does not take into account racial diversity. International collaborative studies are warranted to examine the clinical significance of anti-NVL Abs in other ethnicities.

## Data Availability

All data produced in the present study are available upon reasonable request to the authors.

## Abbreviations

Ab: antibody
ACA: anti-centromere antibody
ANA: anti-nuclear antibody
ARA: anti-RNA polymerase III antibody
ATA: anti-topoisomerase I antibody
DIMT1: DIM1 rRNA methyltransferase ribosome maturation factor
DM: dermatomyositis
ELISA: enzyme-linked immunosorbent assay
HC: healthy control
NVL: nuclear valosin-containing protein-like
IIF: indirect immunofluorescence
ILD: interstitial lung disease
IP: immunoprecipitation
IP-MS: immunoprecipitation combined with gel-free liquid chromatography-tandem mass spectrometry
IPO: intestinal pseudo-obstruction
NUC-ANA: nucleolar patterned anti-nuclear antibody
PH: pulmonary hypertension
PVDF: polyvinylidene difluoride
PWAS: proteome-wide autoantibody screening
SDS-PAGE: sodium dodecyl sulfate-polyacrylamide gel electrophoresis
SLE: systemic lupus erythematosus
SSc: systemic sclerosis
TERF1: telomeric repeat binding factor 1
WPA: wet protein arrays

## Acknowledgements

We appreciate Dr. Kotaro Tsuboyama, MD, PhD, the Lecturer at Biomolecular Design Engineering Laboratory, Institute of Industrial Science, The University of Tokyo, for his advice on our epitope analysis. We thank Ms. Maiko Enomoto, Ms. Teruko Tani, Ms. Mayumi Odagiri, Ms. Yoko Sato, and Ms. Natsuho Yoshifuji for their technical assistance and secretary work.

## Funding sources

None.

## Conflict-of-interest statement

K Yamaguchi, T Okumura, C Ono, K Ogawa, A Miya, A Sato, and N Goshima were employed by ProteoBridge Corporation. T Fukasawa and A Yoshizaki belong to the Social Cooperation Program, Department of Clinical Cannabinoid Research, The University of Tokyo Graduate School of Medicine, Tokyo, Japan, supported by Japan Cosmetic Association and Japan Federation of Medium and Small Enterprise Organizations. T Okamura belongs to the Social Cooperation Program, Department of Functional Genomics and Immunological Diseases, The University of Tokyo Graduate School of Medicine, Tokyo, Japan, supported by Chugai Pharmaceutical Corporation. The remaining authors declare that the research was conducted in the absence of any commercial or financial relationships that could be construed as a potential conflict of interest.

## Author Contributions

KM Matsuda primarily engaged in sample collection in the Tokyo cohort, gathering clinical information, autoantibody measurement, data analysis, and writing the first draft of the manuscript. H Kotani was in charge of epitope mapping. T Matsushita gathered samples and clinical information of the Kanazawa cohort, and also was responsible for IP assays. K Yamaguchi, T Okumura, C Ono, K Ogawa, A Miya, A Sato, and N Goshima prepared wet protein arrays and participated in data analysis. R Uchino, Y Murakami and H Matsunaka provided technical assistance for autoantibody measurement. and sample collection. M Kono, T Okamura, H Shoda and K Fujio participated in sample collection of SLE. T Hisamoto, R Kawanabe, Y Norimatsu, A Kuzumi, T Fukasawa, and A Yoshizaki-Ogawa played roles in clinical information acquisition of SSc and DM. S Sato conceptualized the study. A Yoshizaki conceptualized, launched, and supervised this study, and was involved in revising the manuscript.

---

**Supplementary Figure 1. Serum levels of two other candidate autoantibodies.** Serum levels of anti-DIMT1 and anti-TERF1 Abs in the sera of SSc patients with NUC-ANAs of specific autoantibody unknown (Unknown), ATA positive SSc patients, ACA positive SSc patients, ARA positive SSc patients, DM patients, SLE patients, and HC.

## Notes

### Competing Interest Statement

K Yamaguchi, T Okumura, C Ono, K Ogawa, and N Goshima were employed by ProteoBridge Corporation. T Fukasawa and A Yoshizaki belong to the Social Cooperation Program, Department of Clinical Cannabinoid Research, The University of Tokyo Graduate School of Medicine, Tokyo, Japan, supported by Japan Cosmetic Association and Japan Federation of Medium and Small Enterprise Organizations. T Okamura belongs to the Social Cooperation Program, Department of Functional Genomics and Immunological Diseases, The University of Tokyo Graduate School of Medicine, Tokyo, Japan, supported by Chugai Pharmaceutical Corporation.

### Funding Statement

This study did not receive any funding.

### Author Declarations

This study has been approved by The University of Tokyo Ethical Committee (Approval number 0695).

