## Supplementary figures and images for "Autoantibodies to nuclear valosin-containing protein-like protein: identification and characterization of systemic sclerosis-related anti-nucleolar antibodies utilizing *in vitro* human proteome"

### Supplementary Figure 1

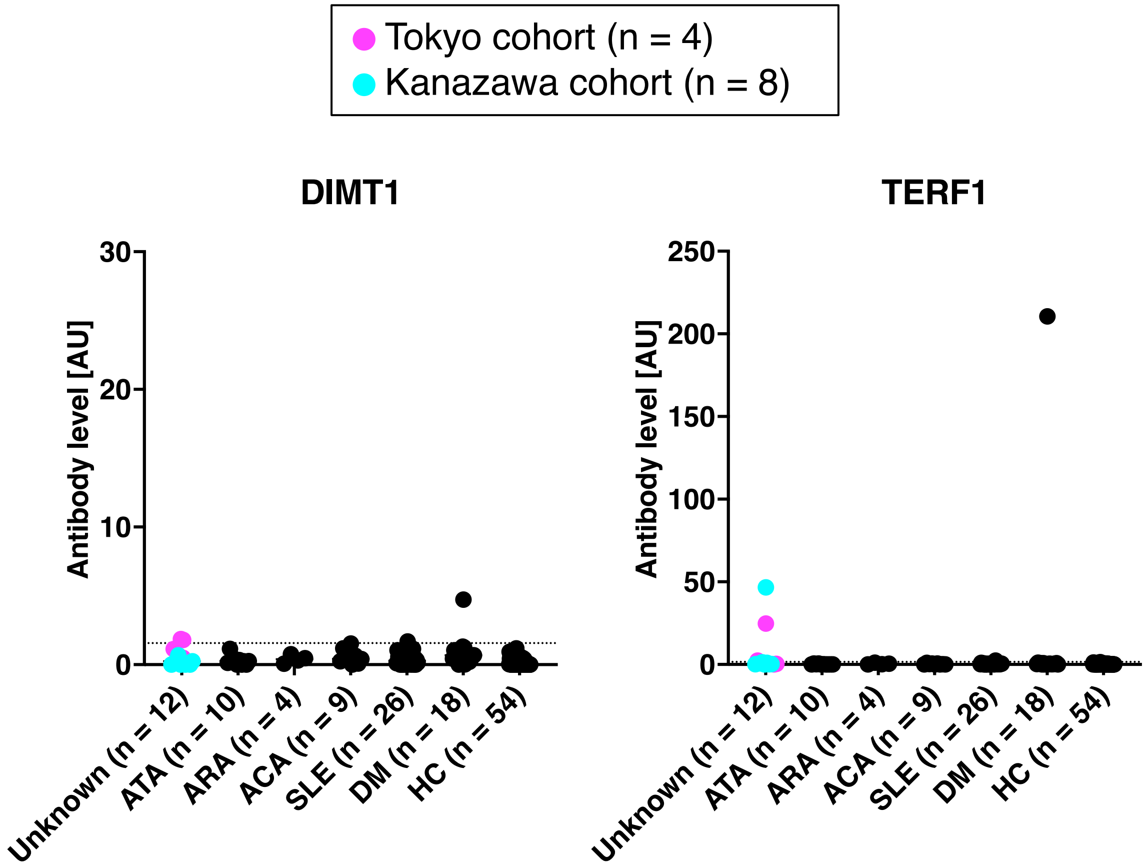


Matsuda KM et al.
Figure S1
